# Multi-resolution vision transformer model for skin cancer subtype classification using histopathology slides

**DOI:** 10.1101/2025.01.28.25321234

**Authors:** Abadh K Chaurasia, Patrick W Toohey, Helen C Harris, Alex W Hewitt

## Abstract

**Introduction:** Digital pathology has significantly advanced cancer diagnosis by enabling high-resolution visualisation and assessment of tissue specimens. However, the manual analysis of these images remains labour-intensive and susceptible to human error, resulting in inconsistencies in diagnosis and treatment decisions. Herein, we developed and externally validated a multi-resolution model for classifying subtypes of skin cancer from whole slide images ( WSIs).

**Methods:** We constructed a dataset comprising approximately 1.13 million histological patches (dividing the WSIs into non-overlapping tiles) from the Non-Melanoma Skin Cancer Segmentation (NMSCS) and Heidelberg datasets. All the patches were normalised using the Macenko method before training a self-supervised vision transformer-based model to classify the most common subtypes of skin cancer: basal cell carcinoma (BCC), squamous cell carcinoma (SCC), intraepidermal carcinoma (IEC), Melanoma, Naevi, and Non-cancerous. Our multi-resolution model was designed to classify melanoma and non-melanoma skin cancer subtypes, incorporating multi-resolution data at 10x, 20x, 40x, and 400x magnifications. The model was externally validated on 5,147 slides from 4,066 patients for non-melanoma cancer subtypes. The model’s performance was evaluated using classification metrics, and the quadratic weighted Cohen’s Kappa (*κ*) score was used to measure the agreement between the model’s predictions and the actual labels with a 95% confidence interval (CI).

**Results:** Our multi-resolution model demonstrated strong classification performance across six classes, achieving an overall *κ* score of 0.859 (95% CI: 0.851, 0.866) and 0.898 (95% CI: 0.892, 0.904) on the validation and testing sets, respectively, reflecting robust performance across diverse skin cancer subtypes. The multi-resolution model for non-melanoma skin cancer exhibited superior performance, achieving an overall *κ* score of 0.919 (95% CI: 0.914, 0.924) on the validation set. On the testing set, the *κ* score ranged from 0.996 to 0.889 with magnifications of 10x, 20x, 40x, and 400x. The attention maps highlighted clinically relevant features for cancerous tissue at different magnifications. Additionally, the model obtained a *κ* score of 0.791 (95% CI: 0.774, 0.808) on the external data at the slide level, indicating substantial agreement between the model’s prediction and the actual label of WSIs.

**Conclusion:** Our multi-resolution model has the potential to assist anatomical pathologists in automatically detecting, highlighting, and classifying subtypes of melanoma and non-melanoma skin cancer subtypes directly from WSIs. This capability could ultimately improve patient outcomes and more effective clinical decision-making in digital pathology. For non-melanoma cancer, our model could be deployed in regions with limited access to experienced dermatopathologists and a high incidence of the disease, particularly in low-resource settings.

## INTRODUCTION

Skin cancer, including melanoma and non-melanoma, is one of the most prevalent forms of malignancy worldwide.^1,2,3^ The incidence is rapidly increasing across diverse populations, especially in Caucasians, leading to a substantial public health burden.^2,3^ The spectrum of skin cancers varies widely, grading from relatively slow-growing cancer, basal cell carcinoma (BCC), to more aggressive forms of cancer such as melanoma. Non-melanoma—BCC, squamous cell carcinoma (SCC)—is the most frequent and generally less deathly, with a lower mortality rate, whereas melanoma, though less common, is highly aggressive with rapid progression and likely high metastatic and accounts for the majority of skin cancer-related mortality.^1,2,4^ Currently, the burden of melanoma is estimated around 0.3 million new cases and 57,000 deaths globally, with the highest incidence rates observed in Australia and New Zealand, and if present trends continue, melanoma cases could increase by around 50% and deaths by 68% by 2040.^1^ In 2022, Australia had the highest global incidence of skin cancer, with Denmark ranking second.^5^ Over two-thirds of Australians may need at least one excision for skin cancers in their lifetime, and deaths from these cancers have nearly doubled in the past two decades.^6,7^ The financial impact of skin cancer is substantial^8,9,10^, emphasising the critical need for effective diagnosing and screening strategies, particularly in high-risk regions like Australia, where UV radiation levels are markedly elevated.^9^

Histopathological examination of skin biopsies remains the gold standard, as pathologists analyse tissue samples using a microscope to identify malignant tissue and determine the cancer subtypes from whole slide images (WSIs). However, it is labour-intensive, time-consuming, and subject to inter-observer variability, leading to inconsistencies in diagnosis and treatment decisions. Modern pathology has significantly transformed anatomical pathology, shifting from traditional microscope-based examination to digital analysis on computer screens. With digital tools, pathologists can directly annotate, measure, and highlight specific areas of interest in the image, making the diagnostic process more streamlined. Despite advancements in digital scanning, current diagnostic methods still require improvement to ensure accurate and consistent diagnoses and efficiently manage the pathological workload in general clinical practices. Integrating digital images and utilising advanced computer vision techniques can transform digital diagnostic pathology, reducing variability, ensuring consistent outcomes, and improving efficiency while reducing workload in traditional histopathology.

AI-driven systems, particularly convolutional neural networks and Vision Transformers (ViT), have demonstrated remarkable accuracy in classifying different types of skin cancers from histopathological images, often surpassing the performance of experienced pathologists.^11,12,13,14^ Most existing AI models have trained on limited and homogenous datasets that target specific cancer subtypes, which often reduces their generalisability and clinical utility for underrepresented subtypes in diverse populations.^15,16,17,18,19^ The need for annotated data for specific subtypes often exacerbates this issue, leading to their exclusion from the analysis. Consequently, the model’s robustness and applicability across diverse clinical scenarios may be compromised, reducing its effectiveness in real-world settings where a more comprehensive range of subtypes is seen. Additionally, using diverse scanners with varying magnifications introduces further variability, complicating the model’s ability to generalise across different imaging conditions. These challenges underscore the importance of incorporating a more comprehensive set of subtypes and developing a robust approach that can comprehensively handle variability introduced by different scanners and magnifications of histological slides, enhancing the model’s generalisability and clinical applicability.

Most AI models for skin cancer classification currently rely on single-resolution data, which may miss the full complexity of histopathological features. This study presents a multi-resolution model for classifying comprehensive subtypes of skin cancer, including melanoma and non-melanoma. We utilise a self-supervised-based ViT model with the Distillation with No Labels (DINOv2) method to detect and classify various skin cancer subtypes using hematoxylin and eosin (H&E)-stained WSIs of skin biopsies. This work allows for detecting and highlighting different subtypes of skin cancer tissues directly from WSIs, assisting pathologists in making accurate diagnoses and enhancing clinical practices efficiently. Our approach can potentially revolutionise modern pathology, improving diagnostic accuracy and reproducibility, especially in areas with limited access to experienced pathologists, such as remote or underserved populations.

## METHODS

### Heidelberg Dataset

The Heidelberg Dataset (HD), a collaborative effort, was initially extracted from the archives of the Institute of Pathology at Heidelberg University, the MVZ Histology, Cytology and Molecular Diagnostics Trier, and the Institute for Dermatopathology, Hannover.^20^ All slides were scanned using an automated slide scanner (Aperio AT2, Leica Biosystems, Nussloch, Germany) with 400x magnification—publicly available.^21^

The histopathological patches were extensively annotated for 16 categories from 386 cases by Frithjof Lobers and Mark Kriegsmann using the QuPath annotation tool.^22^ The categories included chondral tissue, dermis, elastosis, epidermis, hair follicle, skeletal muscle, necrosis, nerves, sebaceous glands, subcutis, eccrine glands (sweat glands), vessels, BCC, SCC, naevi, and melanoma. All the patches from the slide were generated with 100 x 100 µm (∼395 x 395 px) in size using QuPath, ensuring the precision and thoroughness of the data. The patients’ slides were divided into training, validation, and test sets, with each patient’s patches assigned to only one set. These split data were unchanged throughout our study, as shown in **eTable S1** in the supplemental information.

### Non-melanoma Skin Cancer Segmentation Dataset

The Non-Melanoma Skin Cancer Segmentation (NMSCS) dataset includes 290 H&E slides provided by MyLab Pathology.^23^ This diverse dataset includes 140 BCC, 60 SCC, and 90 intraepidermal carcinoma (IEC) cases, covering over 90% of skin cancer subtypes.^24^ The dataset includes 100 shave, 58 punch, and 132 excision biopsies from patients aged 34 to 96 with a median age of 70, ensuring a broad age range representation. A pathologist meticulously hand-annotated each slide to indicate which tissue section most represented the cancer class, ensuring the accuracy and reliability of the dataset. The slides were scanned using a DP27 Olympus microscope camera at 10x magnification. The pixel width corresponds to 0.67µm, resulting in images ranging from 11 million to 500 million pixels.

This dataset includes downsampled versions (1x, 2x, 5x, and 10x) with slide images and corresponding 12 class segmentation mask colours. The ground-truth segmentations were created in ImageJ, assigning each pixel to one of 12 categories: Glands, Inflammation, Hair Follicles, Hypodermis, Reticular Dermis, Papillary Dermis, Epidermis, Keratin, Background, BCC, SCC, and IEC.^23^ Notably, the ideal healthy epidermis was labelled as Epidermis, while dysplastic keratinocytes and carcinoma features were included in the IEC class to address the variation within the epidermis.

For our study, the patches were generated at three magnification levels (10x, 20x, and 40x) from 10x slides to establish a mapping between RGB colour codes and tissue classes. We resized the images and masks using the Lanczos filter and nearest-neighbour interpolation to simulate different magnification levels.^25^ We then divided the rescaled images into 518 x 518 pixels without overlapping patches. To ensure the quality and relevance of the patches, we applied a tissue content threshold, discarding patches with less than 20% tissue content in a patch. Additionally, we implemented a classification criterion requiring each patch to contain a minimum of 80% of a specific tissue type to be retained for analysis. Each retained patch was categorised according to its predominant tissue type based on predefined colour mappings.^23^ This approach allowed us to generate comprehensive patches across multiple resolutions. Finally, we categorise four classes: BCC, SCC, IEC, and Non-cancerous, as presented in **Table 1**.

**Table 1.** Distribution of cancerous and non-cancerous patches from the Heidelberg and Non-melanoma Skin Cancer Segmentation Dataset.

| Dataset type | Total patches | Dataset name | Number of slides | Magnification | Number of BCC patches | Number of SCC patches | Number of Melanoma patches | Number of Naevi patches | Number of IEC patches | Number of Non-cancerous patches |
| --- | --- | --- | --- | --- | --- | --- | --- | --- | --- | --- |
| Training | 875277 | NMSCS | 231 | 10x | 592 | 1801 | - | - | 279 | 26097 |
|  |  |  |  | 20x | 3318 | 8650 | - | - | 2269 | 124351 |
|  |  |  |  | 40x | 16197 | 38331 | - | - | 12882 | 551539 |
|  |  | HD | - | 400x | 6919 | 6793 | 7784 | 7923 | - | 59552 |
| Validation | 90553 | NMSCS | 28 | 10x | 66 | 225 | - | - | 11 | 2451 |
|  |  |  |  | 20x | 359 | 1109 | - | - | 189 | 11947 |
|  |  |  |  | 40x | 1748 | 5099 | - | - | 1221 | 53774 |
|  |  | HD | - | 400x | 1063 | 919 | 1220 | 944 | - | 8208 |
| Testing | 161088 | NMSCS | 31 | 10x | 41 | 175 | - | - | 22 | 4679 |
|  |  |  |  | 20x | 251 | 840 | - | - | 255 | 22253 |
|  |  |  |  | 40x | 1172 | 3690 | - | - | 1573 | 98098 |
|  |  | HD | - | 400x | 941 | 3470 | 2678 | 1762 | - | 19188 |

Abbreviations: **NMSCS** = Non-Melanoma Skin Cancer Segmentation, **HD** = Heidelberg Dataset, **BCC** = Basal Cell Carcinoma, **SCC** = Squamous Cell Carcinoma, **IEC** = Intraepidermal Carcinoma.

### COBRA dataset

The COBRA (Classification Of Basal cell carcinoma, Risky skin cancers and Abnormalities) dataset was collected from the Radboud University Medical Center and scanned using a 3DHistech Pannoramic 1000 scanner at 20x magnification (0.24 μm).^15^ This dataset comprises 5,147 slides from 4,066 patients, with approximately half having BCC tumours (2,673) and the rest exhibiting non-BCC slides (2,474), as confirmed by the pathology reports. The pathology findings revealed that 24% of slides depicted superficial BCC, 69% showed nodular BCC, 24% displayed micro-nodular BCC, and 33% exhibited infiltrative BCC. Notably, some slides showed multiple types of BCC, indicating a diverse presentation in the samples. This dataset is publicly available.^26^

### Image Processing and Patch Extraction

Before training the model, a comprehensive preprocessing pipeline was implemented to standardise the input patches (518 x 518) for model training. The patches were first normalised using the Macenko method using the staintools library to reduce staining variability across histopathological slides.^27,28^ This involved fitting a target image to the StainNormalizer and applying this normalisation to all H&E-stained patches. We also implemented data augmentation techniques such as horizontal and vertical flipping, rotation, and lighting adjustments to enhance the dataset’s variability while preserving tissue characteristics; parameters for data augmentation and transformation techniques are applied in **eTable S2** in supplemental information.

### Model Architecture and Training Procedures

The ViT model with registers (capturing complex visual representations) was selected for the classification task^29^, offering a novel approach compared to traditional convolutional neural networks.^30^ The ViT model uses a patch-based approach to divide the input image into 14 x 14 smaller patches, which are then transformed into high-dimensional vectors. The Transformer uses multi-head self-attention mechanisms to process these vectors, initially designed for natural language tasks. The self-attention mechanism allows the model to weigh the importance of each patch relative to others, capturing global context and dependencies across the entire image. These features enable the model to capture long-range dependencies within the image, making it particularly suitable for analysing complex histopathological images. The model, vit_base_patch14_reg4_dinov2.lvd142m, was initially trained on a diverse dataset of 142 million visual representations using the DINOv2, a self-supervised learning approach.^31,32^ DINOv2 allows the model to learn from unlabeled data by creating pseudo-labels through a teacher-student training framework. The teacher network, typically more stable and accurate, generates targets for the student network to learn, resulting in a robust and generalisable feature representation.^31^

We employed the Fastai framework to implement a transfer learning approach, starting with pre-trained weights.^33,34^ The model was fine-tuned for ten epochs using a learning rate of 1.2e-3. A callback function was implemented to monitor validation loss with a patience of 3 and a minimum delta of 0.01, along with a weight decay of 3e-3.^35^ After fine-tuning, the model was subsequently unfrozen, allowing for training for an additional ten epochs using a 1-cycle policy, with a learning rate slice ranging from 1e-6 to 1e-4. This phase also incorporated the same callbacks for early stopping and model saving, with weight decay adjusted to 2e-3. The 1-cycle policy, known for its efficacy in deep learning, dynamically adjusted the learning rate throughout training, starting with a low rate, peaking, and then decreasing to avoid suboptimal local minima and accelerate convergence.^35,36^

This study implemented a multi-resolution model to evaluate their effectiveness in detecting cancerous tissue at patch and biopsy levels (WSIs). The multi-resolution model was trained on the most prevalent subtypes of skin cancer from two different datasets with a magnification of 10x, 20x, 40x, and 400x (**Table 1**) to classify broad subtypes of skin cancer: BCC, SCC, IEC, Melanoma, Naevi, and Non-cancerous. The data for melanoma and naevi was only available at a single resolution of 400x for training. Therefore, we further refined the classes that only contained common types of cancerous class from both datasets to develop a final robust multi-resolution model for classifying the non-melanoma subtype of skin cancer. This step triggered the model to capture complex features from a diverse population at various magnifications.

### Attention Maps Visualization

The attention maps enhance the model’s transparency for understanding its decision-making process in a clinical setting. To enhance the clinical interpretability of the model’s predictions, attention maps were generated to emphasise the areas of the input images that were most significant in the model’s classification decisions. The patches were preprocessed by resizing them to 518 x 518 pixels and padding them to ensure they were compatible with the model’s input patch size. The attention maps were extracted from all attention heads (12) in the final self-attention layers of the model, where the attention weights were aggregated and processed to highlight the model’s focus on specific regions in images. These heatmaps were normalised and overlapped onto the original images, providing visual insight into the focus areas during the model’s decision-making process. The model’s predicted class and associated confidence scores were displayed alongside each image to contextualise the attention map further.

### Slide-Level Cancer Detection

Cancer detection at the slide level was determined by dividing the WSIs into non-overlapping patches, each sized 518 x 518 pixels. Patches then underwent preprocessing to filter out those with insufficient tissue content, applying thresholds for tissue presence (20%) and excluding predominantly white or black patches. The trained model was then used to predict each patch, identifying the subtypes of cancerous tissue. Consequently, the slide-level prediction was determined by aggregating the patch predictions from a slide (WSIs). If more than 5% of the total patches were classified as cancerous patches, we considered that slide as cancerous, and next, the slide was labelled according to the most frequent cancerous class; otherwise, it was classified as a Non-cancerous class.

Additionally, visualisations were created to highlight the regions of the WSIs identified as cancerous and Non-cancerous tissue within slides. These visualisations involved overlaying colour-coded rectangles on the original slide image based on the predicted class and creating high magnification of the detected class patch to provide a detailed view of the cancerous and Non-cancerous tissue.

### Model Evaluation and Statistical Analysis

The model’s performance was assessed against the ground truth labels using a comprehensive set of classification metrics, including accuracy, precision, recall, F1-score, quadratic weighted Cohen’s Kappa (*κ*) score, and the area under the receiver operating characteristic curve (AUROC). All the metrics were calculated for each class against all others (one-vs-rest classes).^37^ To strengthen the robustness of the evaluation, a bootstrap resampling method was applied^38^, generating 4,000 bootstrap samples from the testing datasets. Each sample was used to recalculate the metrics, allowing for the estimation of the mean value and the 95% confidence interval (CI) for each metric.

All experiments were conducted on a virtual Ubuntu desktop (version 22.04), utilising an NVIDIA A100 GPU and 40GB of RAM. The statistical analyses and models training were performed using Python, Fastai, and PyTorch libraries.^39,40,41^

## RESULTS

### Performance of Multi-Resolution Model

Our multi-resolution model demonstrated strong classification performance across six classes: BCC, IEC, Melanoma, Naevi, SCC, and Non-cancerous. On the validation set, it obtained an overall *κ* score of 0.859 (95% CI: 0.851, 0.866), indicating its effectiveness in distinguishing between different subtypes of skin cancer, with an AUROC exceeding 99% for all classes, as shown in **eTables S3** in the supplemental information. Additionally, the model achieved an overall *κ* score of 0.898 (95% CI: 0.892, 0.904) on the testing sets, indicating strong performance across various skin cancer subtypes. This performance is illustrated in **Table 2**, and the predicted patches with their actual labels can be seen in **eFigure S1** in the supplemental information. The confusing matrixes highlighted in **eFigures S2 and S3** in the supplemental information indicate that the melanoma class achieved an accuracy of over 93% on both validation and testing sets. However, the IEC class achieved only 64.04% on the validation and 70.86% on the testing set. The top 20 misclassified patches were visualised in **eFigure S4** of the supplemental information. Our final multi-resolution model for classifying non-melanoma skin cancer subtypes achieved an overall *κ* score of 0.919 (0.914, 0.924) on the validation set, with each resolution contributing to its ability to capture features at different scales. The model was also evaluated on the testing set at various resolutions, indicating high classification accuracies across all the resolutions, summarised in **Table 3**.

**Table 2.**
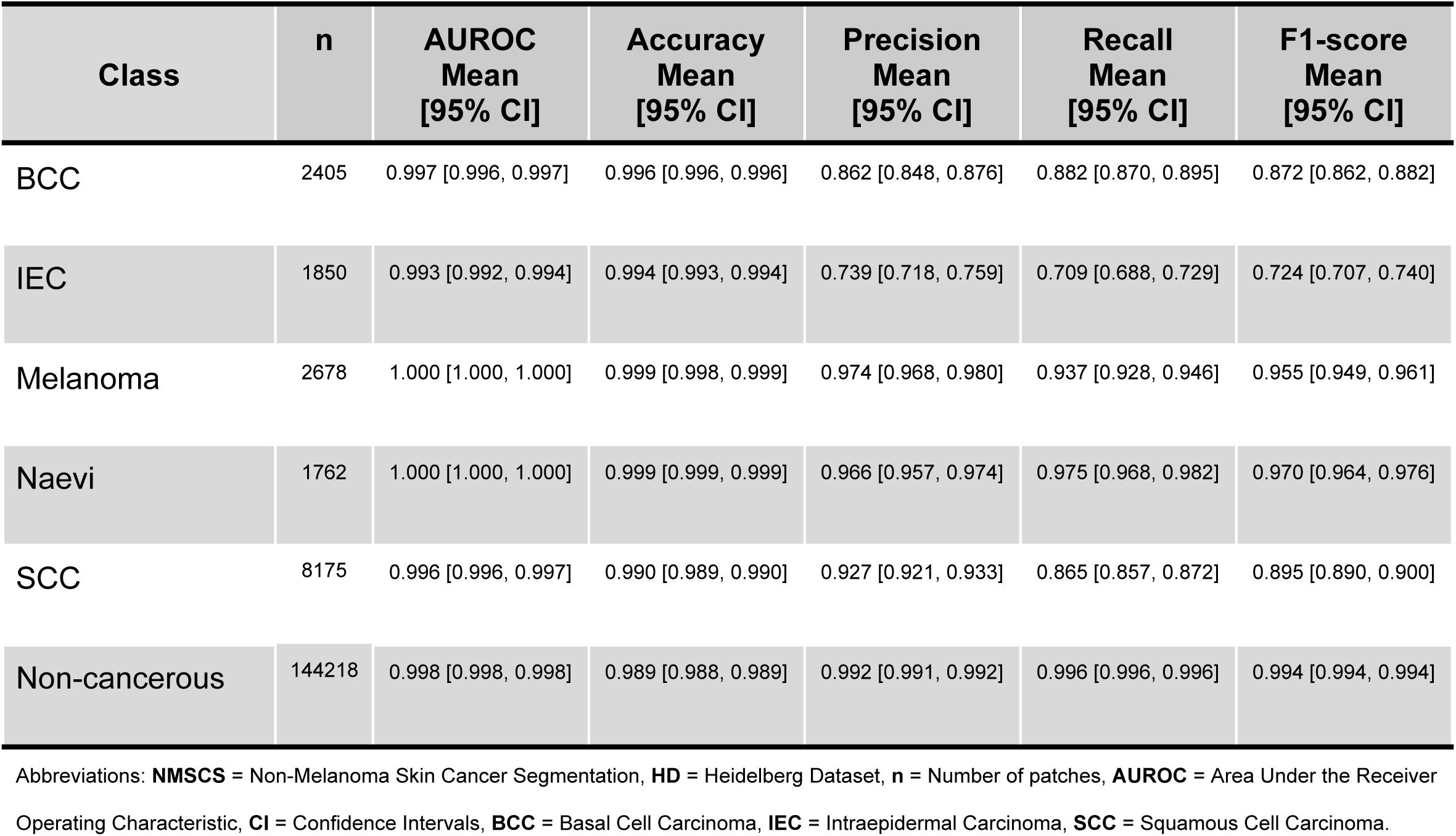
Performance of the *multi-resolution model on testing sets, 161,088 patches at 10x, 20x, 40x, and 400x magnifications from* NMSCS and HD *datasets*.

| Class | n | AUROC Mean [95% CI] | Accuracy Mean [95% CI] | Precision Mean [95% CI] | Recall Mean [95% CI] | F1-score Mean [95% CI] |
| --- | --- | --- | --- | --- | --- | --- |
| BCC | 2405 | 0.997 [0.996, 0.997] | 0.996 [0.996, 0.996] | 0.862 [0.848, 0.876] | 0.882 [0.870, 0.895] | 0.872 [0.862, 0.882] |
| IEC | 1850 | 0.993 [0.992, 0.994] | 0.994 [0.993, 0.994] | 0.739 [0.718, 0.759] | 0.709 [0.688, 0.729] | 0.724 [0.707, 0.740] |
| Melanoma | 2678 | 1.000 [1.000, 1.000] | 0.999 [0.998, 0.999] | 0.974 [0.968, 0.980] | 0.937 [0.928, 0.946] | 0.955 [0.949, 0.961] |
| Naevi | 1762 | 1.000 [1.000, 1.000] | 0.999 [0.999, 0.999] | 0.966 [0.957, 0.974] | 0.975 [0.968, 0.982] | 0.970 [0.964, 0.976] |
| SCC | 8175 | 0.996 [0.996, 0.997] | 0.990 [0.989, 0.990] | 0.927 [0.921, 0.933] | 0.865 [0.857, 0.872] | 0.895 [0.890, 0.900] |
| Non-cancerous | 144218 | 0.998 [0.998, 0.998] | 0.989 [0.988, 0.989] | 0.992 [0.991, 0.992] | 0.996 [0.996, 0.996] | 0.994 [0.994, 0.994] |

Abbreviations: **NMSCS** = Non-Melanoma Skin Cancer Segmentation, **HD** = Heidelberg Dataset, **n** = Number of patches, **AUROC** = Area Under the Receiver Operating Characteristic, **CI** = Confidence Intervals, **BCC** = Basal Cell Carcinoma, **IEC** = Intraepidermal Carcinoma, **SCC** = Squamous Cell Carcinoma.

**Table 3.** Performance of the multi-resolution model for non-melanoma skin cancer subtypes was evaluated on testing sets at different magnifications, presenting the mean along with 95% confidence intervals (CI).

| Metric | Class | 10x<br>(NMSCS) | 20x<br>(NMSCS) | 40x<br>(NMSCS) | 400x<br>(HD) |
| --- | --- | --- | --- | --- | --- |
| **AUROC** | **BCC** | 0.999 [0.998, 1.000] | 0.999 [0.998, 1.000] | 0.997 [0.996, 0.998] | 0.999 [0.998, 1.000] |
|  | **SCC** | 1.000 [1.000, 1.000] | 0.999 [0.999, 0.999] | 0.997 [0.996, 0.997] | 0.998 [0.997, 0.998] |
|  | **Non-cancerous** | 1.000 [1.000, 1.000] | 0.999 [0.999, 0.999] | 0.997 [0.996, 0.997] | 0.999 [0.998, 0.999] |
| **Accuracy** | **BCC** | 1.000 [0.999, 1.000] | 0.999 [0.999, 1.000] | 0.999 [0.998, 0.999] | 0.995 [0.995, 0.996] |
|  | **SCC** | 0.997 [0.996, 0.999] | 0.995 [0.994, 0.996] | 0.992 [0.991, 0.992] | 0.985 [0.983, 0.986] |
|  | **Non-cancerous** | 0.997 [0.996, 0.999] | 0.995 [0.994, 0.996] | 0.991 [0.990, 0.991] | 0.987 [0.986, 0.989] |
| **Precision** | **BCC** | 1.000 [1.000, 1.000] | 0.975 [0.954, 0.992] | 0.952 [0.939, 0.964] | 0.909[0.890, 0.926] |
|  | **SCC** | 0.994 [0.980, 1.000] | 0.946 [0.930, 0.961] | 0.909 [0.900, 0.919] | 0.972 [0.966, 0.978] |
|  | **Non-cancerous** | 0.997 [0.996, 0.999] | 0.996 [0.996, 0.997] | 0.994 [0.993, 0.994] | 0.990 [0.988, 0.991] |
| **Recall** | **BCC** | 0.975 [0.915, 1.000] | 0.952 [0.924, 0.976] | 0.923 [0.908, 0.938] | 0.984 [0.976, 0.992] |
|  | **SCC** | 0.931 [0.890, 0.965] | 0.918 [0.900, 0.936] | 0.851 [0.839, 0.862] | 0.922 [0.913, 0.931] |
|  | **Non-cancerous** | 1.000 [0.999, 1.000] | 0.998 [0.997, 0.998] | 0.996 [0.996, 0.997] | 0.995 [0.994, 0.996] |
| **F1-score** | **BCC** | 0.987 [0.955, 1.000] | 0.964 [0.946, 0.979] | 0.937 [0.927, 0.947] | 0.945 [0.934, 0.955] |
|  | **SCC** | 0.961 [0.938, 0.981] | 0.932 [0.919, 0.944] | 0.879 [0.871, 0.887] | 0.946 [0.941, 0.952] |
|  | **Non-cancerous** | 0.999 [0.998, 0.999] | 0.997 [0.997, 0.998] | 0.995 [0.995, 0.995] | 0.992 [0.991, 0.993] |
| $\kappa$ | **BCC** | 0.966 [0.947, 0.983] | 0.937 [0.925, 0.948] | 0.888 [0.881, 0.895] | 0.922[0.913, 0.932] |
|  | **SCC** | 0.966 [0.947, 0.983] | 0.936 [0.924, 0.947] | 0.888 [0.881, 0.896] | 0.922 [0.912, 0.932] |
|  | **Non-cancerous** | 0.966 [0.948, 0.983] | 0.936 [0.924, 0.947] | 0.888 [0.880, 0.895] | 0.922 [0.913, 0.931] |

Abbreviations: **NMSCS** = Non-Melanoma Skin Cancer Segmentation, **HD** = Heidelberg Dataset, **AUROC** = Area Under the Receiver Operating Characteristic, **BCC** = Basal Cell Carcinoma, **IEC** = Intraepidermal Carcinoma, **SCC** = Squamous Cell Carcinoma, $\kappa$ = Quadratic weighted Cohen's Kappa score.

### Attention Maps for Model Interpretability

The multi-resolution model generated attention maps from the testing sets at different magnifications. The heatmaps indicated areas of higher relevance, enhancing the understanding of how the model distinguishes between cancerous and non-cancerous tissue subtypes. Twelve attention heads individually identified unique aspects of the morphological characteristics of images related to melanoma and non-melanoma skin cancer subtypes, as demonstrated in **eFigures S5 and S6** in the supplemental information. The combined attention maps from all the heads provided valuable insights into the model’s decision-making processes, offering transparency for clinical utility and assisting in understanding how specific features influenced classification outcomes, as depicted in **Figure 1**. Additionally, the attention maps highlighted clinically relevant features for non-melanoma skin cancer subtypes at different magnifications, illustrated in **eFigure S7** in the supplemental information.

**Figure 1.**
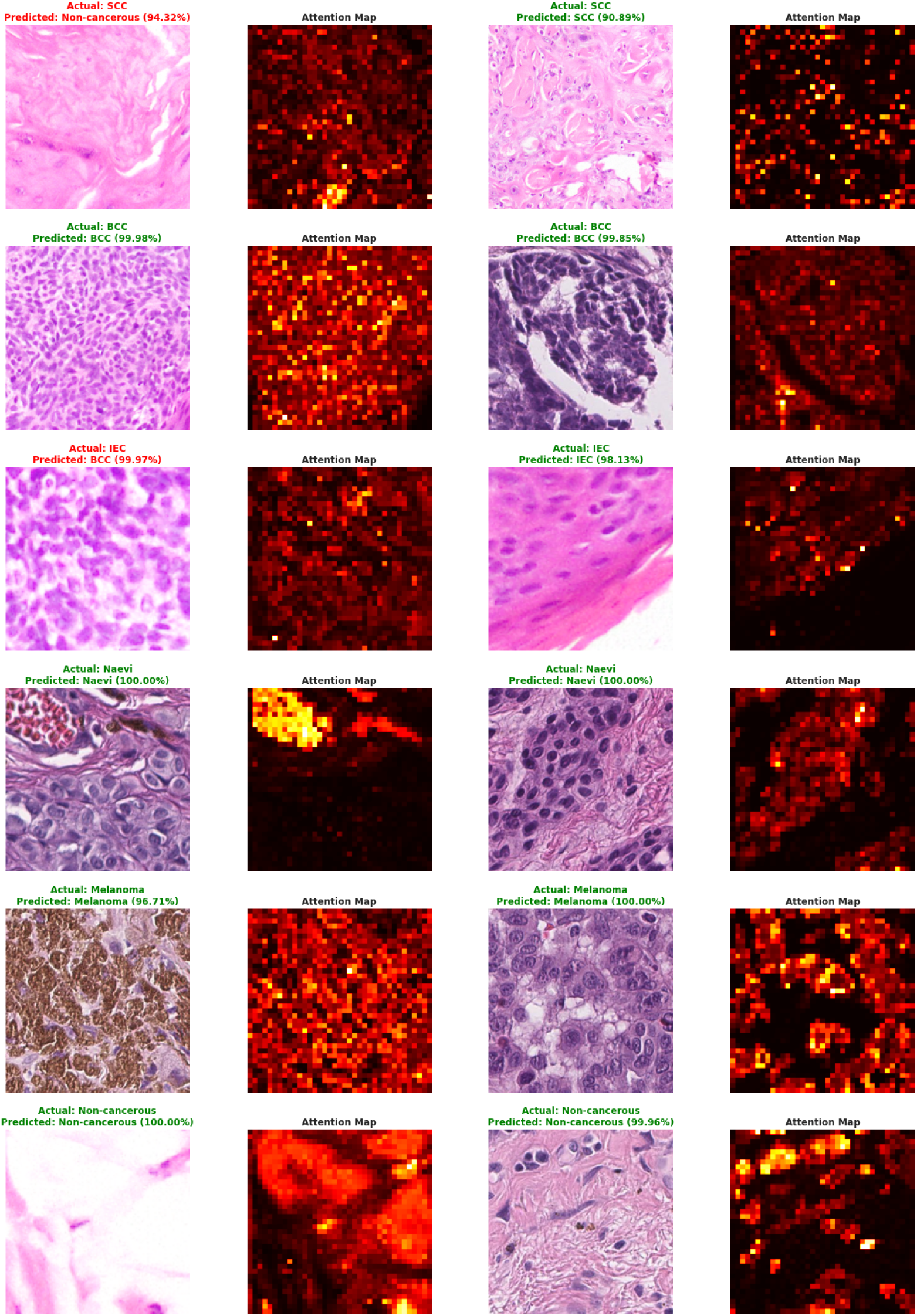
Example attention maps for classifying melanoma and non-melanoma subtypes of skin cancer on the testing sets, showing each pair of patches with attention maps, actual class, and predicted class with prediction probability.

### External Validation and Generalisability

The generalisability and robustness of the multi-resolution models for non-melanoma skin cancer subtype classification were rigorously evaluated on unseen WSIs (5,147 slides) data from the COBRA set, focusing on slide-level detection of BCC versus non-BCC. The model achieved a *κ* score of 0.791 at the native resolution of the slides, with a CI of 0.774 to 0.808, indicating substantial agreement between predicted and actual labels of the slide. Additionally, the model attained an overall accuracy of 0.896 (95% CI: 0.888, 0.904) in classifying subtypes of non-melanoma skin cancer. However, it misclassified 616 non-cancerous slides as SCC and identified 23 slides as BCC. These findings highlight the clinical utility of the multi-resolution model at diverse resolutions of slides, particularly for lower magnifications (20x), which are computationally efficient for deploying the model in digital pathology, as shown in **Table 4**.

**Table 4.**
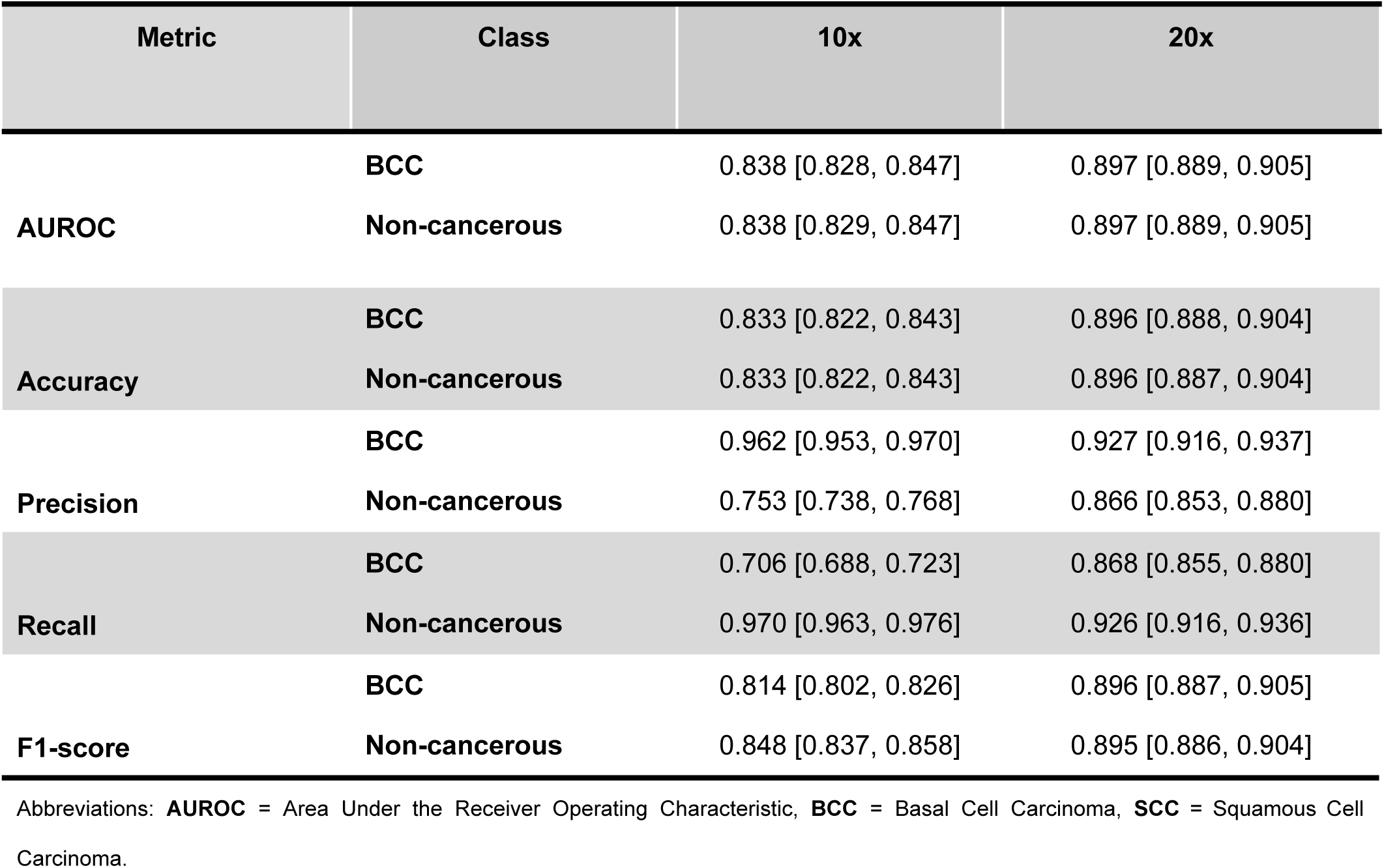
Performance of multi-resolution models for non-melanoma skin cancer subtypes classification at two different magnifications on external data, utilising 2673 BCC and 2474 non-BCC slides from the COBRA dataset.

| Metric | Class | 10x | 20x |
| --- | --- | --- | --- |
| **AUROC** | **BCC** | 0.838 [0.828, 0.847] | 0.897 [0.889, 0.905] |
|  | **Non-cancerous** | 0.838 [0.829, 0.847] | 0.897 [0.889, 0.905] |
| **Accuracy** | **BCC** | 0.833 [0.822, 0.843] | 0.896 [0.888, 0.904] |
|  | **Non-cancerous** | 0.833 [0.822, 0.843] | 0.896 [0.887, 0.904] |
| **Precision** | **BCC** | 0.962 [0.953, 0.970] | 0.927 [0.916, 0.937] |
|  | **Non-cancerous** | 0.753 [0.738, 0.768] | 0.866 [0.853, 0.880] |
| **Recall** | **BCC** | 0.706 [0.688, 0.723] | 0.868 [0.855, 0.880] |
|  | **Non-cancerous** | 0.970 [0.963, 0.976] | 0.926 [0.916, 0.936] |
| **F1-score** | **BCC** | 0.814 [0.802, 0.826] | 0.896 [0.887, 0.905] |
|  | **Non-cancerous** | 0.848 [0.837, 0.858] | 0.895 [0.886, 0.904] |

Abbreviations: **AUROC** = Area Under the Receiver Operating Characteristic, **BCC** = Basal Cell Carcinoma, **SCC** = Squamous Cell Carcinoma.

Furthermore, the multi-resolution model was used to detect and highlight the cancerous tissue within WSIs, providing insights into the relevant regions for emphasising these tissues. **Figure 2** visualises the four slides randomly selected from the COBRA dataset to identify and classify cancerous tissue subtypes, maintaining the original resolution of the slides. The highlighted regions emphasise areas identified as cancerous and Non-cancerous tissues, while the high-magnification patches provide an in-depth visual analysis of the cellular morphology of the detected tissue subtype for assisting pathologists in accurately and efficiently making decisions in digital pathology.

**Figure 2.**
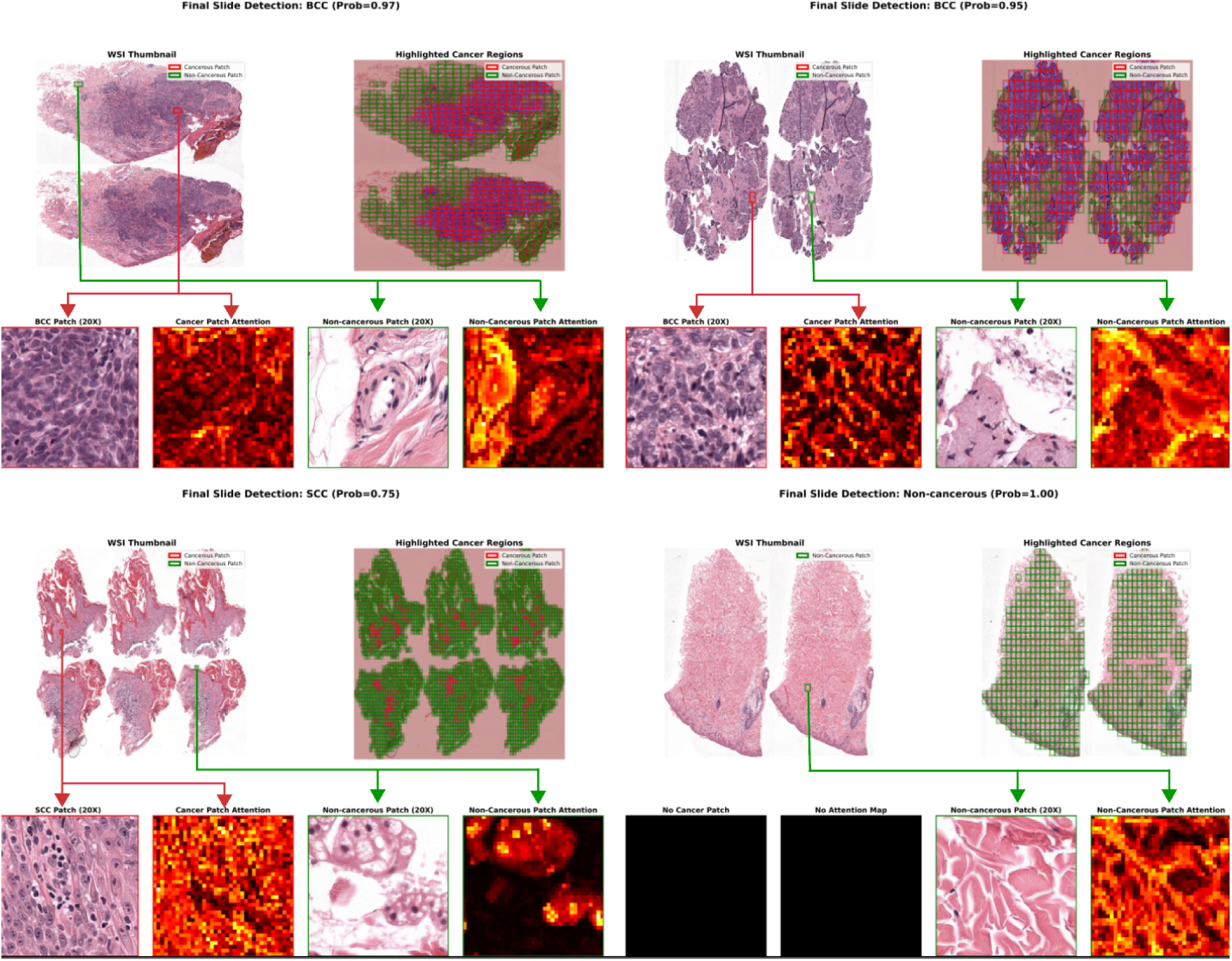
The COBRA dataset was used to classify non-melanoma skin cancer subtypes (BCC, SCC, and Non-cancerous) at the slide level detection. The thumbnails of WSI and highlighted cancer regions indicate cancerous (red) and non-cancerous (green) boxes. The high-magnification patches (20X) were taken from the WSI thumbnail, and their corresponding attention maps are visualised.

## DISCUSSION

Histopathological diagnosis of skin cancer is highly dependent on the expertise of pathologists, leading to variability in interpretations, especially in complex cases or when dealing with rare subtypes of skin cancer. This variability can result in misdiagnosis or delayed diagnosis, impacting patient outcomes. While considerable progress has been made in developing AI models for skin cancer classification, many of these models focus on specific subtypes of skin cancer, such as melanoma, and may not be comprehensive enough to handle the full spectrum of skin cancer subtypes. This study introduced a novel approach for detecting a broad range of subtypes of skin cancer at the histology patch and slide levels using a self-supervised ViT-based model employing the DINOv2 approach with multi-resolution data. Our multi-resolution model accurately identifies different subtypes of skin cancer, even with diverse magnifications and scanner conditions. The model can accurately distinguish between six skin cancer classes, including non-cancerous tissues. It demonstrates high performance across various resolutions on testing sets, highlighting its potential for broader clinical applications in digital pathology.

Although the data for melanoma and naevi were only available at a single resolution of 400x for training, the multi-resolution model demonstrated over 99.9% accuracy on testing sets for melanoma and naevi. The results in **eFigures S2 and S3** in the supplemental information indicate that the melanoma class achieved an accuracy of over 93%, as exemplified by the confusion matrixes. However, the IEC class achieved only 64.04% on the validation and 70.86% on the testing set. This was due to highly imbalanced data; the class with limited data underperformed compared to classes with many patches, even with class weights applied during the training. This imbalance likely impacted the model’s generalisation ability to external datasets or different clinical environments, as evidenced by lower accuracy in the IEC class, and most of the dataset comprises Non-cancerous cases. Therefore, we dropped the melanoma, Naevi, and IEC classes to enhance a robust multi-resolution model for non-melanoma skin cancer classification.

The study by Ianni et al. focused on classifying four types of skin cancer with an overall 78% accuracy on an uncurated test dataset of 13,537 WSIs from multiple sites.^42^ In contrast, our study expanded the scope to include five significant cancer subclasses and one Non-cancerous class, achieving over 99% accuracy across all six classes on the testing set (**Table 2**) and obtaining an overall *κ* score of 0.898 (95% CI: 0.892, 0.904). However, direct comparison could not be feasible due to the different approaches and applied training procedures. This comparison only highlights the scalability and robustness of multi-resolution models, demonstrating their capability to handle a broader range of skin cancer types and slide magnification while maintaining high accuracy in real-world clinical settings. The multi-resolution model for non-melanoma subtype cancer can detect BCC, SCC, and Non-cancerous patches (**Figure 2**) with high AUROC using ViT-based self-supervised learning with DINOv2. The study by Geijs et al. focused on detecting and subtyping BCC using weakly supervised learning.^15^ The best-performing StreamingCLAM model had an AUC of 0.958 from 397 slides for BCC detection, while our model achieved an AUROC of 0.897 across 5,147 WSIs. Our model was externally validated using over 12 times than Geijs’s data.^15^ These findings demonstrate the efficacy of employing advanced techniques in a multi-resolution model, allowing for accurate and efficient classification of non-melanoma subtypes of skin cancer directly from WSIs. The model can be beneficial in low-resource settings with limited access to experienced anatomical dermatopathologists. Thus, the AI-driven solution can potentially bridge this gap by offering consistent and accurate diagnostic support in modern pathology.

This study has some limitations. Firstly, the multi-resolution model for melanoma has yet to be validated on unseen data, which constrains its applicability and generalisability to a broader spectrum of skin cancer types, including Naevi and IEC. Furthermore, the training of the multi-resolution model depended on only two datasets, which may compromise its ability to cover a broader range of clinical characteristics, potentially impacting its robustness, particularly among different ethnic populations. Additionally, the model’s performance may be limited by the data quality included in the training, as variations in imaging techniques and sample preparation could introduce bias. Furthermore, this study overlooked potential confounding such as patient demographics and treatment history, which could influence the model’s performance. Lastly, the model was externally validated only for non-melanoma subtypes of skin cancer, specifically BCC versus non-BCC slides, due to the unavailability of publicly accessible labelled data for other cancer subtypes. Future studies should focus on obtaining multi-resolution data from multiple centres to capture the variability inherent in these tissue subtypes and enhance the model’s clinical utility in detecting broader cancer subtypes.

In conclusion, the multi-resolution model offers a valuable tool for anatomical pathologists by enabling the automatic detection, highlighting, and classification of skin cancer subtypes directly from the WSIs. Our model demonstrates high precision in distinguishing between various subtypes of skin cancer, including melanoma and non-melanoma. For non-melanoma skin cancer, our model can heighten the regions with limited access to experienced anatomical dermatopathologists and a high incidence of non-melanoma skin cancer, especially in low-resource settings and areas with a high incidence of non-melanoma skin cancer. Future research should focus on retrieving diverse ethnic data to strengthen the multi-resolution model’s ability to detect various subtypes of skin cancer from WSIs, thereby improving the model’s utility and generalisability for global populations.

## Data Availability

All data are available online.

## ACKNOWLEDGEMENTS

This work was supported by an Australian National Health and Medical Research Council Leadership Award (A.W.H.).

## AUTHOR CONTRIBUTIONS

A.K.C. was responsible for designing the study, selecting and implementing the self-supervised vision transformer (ViT) architecture, training and validating the models, analysing the results, and preparing the manuscript. H.C.H. reviewed all data utilised in the study and provided interpretations of the results in the clinical context. P.W.T. reviewed and edited the manuscript, analysed classification metrics, and contributed to its revisions. A.W.H. supervised the overall study design, reviewed the clinical significance and validation protocols, interpreted the analyses, and provided valuable suggestions for enhancing the manuscript.

## CONFLICT OF INTEREST

A.K.C., P.W.T., and A.W.H. are co-founders of Pandani Solutions Pty Ltd in Australia, which develops AI-driven tools to assist anatomical pathologists.

## Notes

### Author Declarations

https://www.frontiersin.org/journals/oncology/articles/10.3389/fonc.2022.1022967/full https://www.sciencedirect.com/science/article/pii/S2352340921008623 https://aws.amazon.com/marketplace/pp/prodview-h4lwgyj4myxh6

