## Supplemental information for "Multi-resolution vision transformer model for skin cancer subtype classification using histopathology slides"

**eTable S1** *Patches and patient distribution from the Heidelberg Dataset.*

| Class | Training (n) | | Validation (n) | | Test (n) | |
|---|---|---|---|---|---|---|
| | Patches | Patients | Patches | Patients | Patches | Patients |
| Chondral Tissue | 4442 | 9 | 743 | 2 | 1992 | 7 |
| Dermis | 15878 | 134 | 1857 | 20 | 4875 | 39 |
| Elastosis | 136 | 1 | 6 | 1 | 66 | 1 |
| Epidermis | 10419 | 130 | 1086 | 19 | 2613 | 36 |
| Hair Follicle | 1437 | 104 | 250 | 15 | 325 | 27 |
| Skeletal Muscle | 6159 | 47 | 904 | 7 | 669 | 10 |
| Necrosis | 1641 | 24 | 468 | 5 | 924 | 7 |
| Nerves | 1201 | 93 | 219 | 13 | 464 | 30 |
| Sebaceous Glands | 7268 | 94 | 1074 | 13 | 2565 | 30 |
| Subcutis | 7370 | 64 | 1245 | 9 | 3438 | 26 |
| Sweat Glands | 2533 | 94 | 220 | 11 | 818 | 27 |
| Vessels | 1068 | 109 | 136 | 14 | 439 | 31 |
| BCC | 6919 | 71 | 1063 | 12 | 941 | 10 |
| SCC | 6793 | 61 | 919 | 10 | 3470 | 29 |
| Naevi | 7923 | 72 | 944 | 8 | 1762 | 18 |
| Melanoma | 7784 | 59 | 1220 | 9 | 2678 | 19 |

Abbreviations: **n** =Number of patches or patients, **BCC** = Basal Cell Carcinoma, **SCC** = Squamous Cell Carcinoma.

**eTable S2** *Parameters used for data augmentation and transformation techniques.*

| Transformation | Parameter | Value |
|---|---|---|
| Resize | Size | 518 x 518 pixels |
| Random Rotation | Max Rotate | 20 degrees |
| Zoom | Max Zoom | 1.05x |
| Flipping | Horizontally | True |
| | Vertically | True |
| Lighting Adjust | Max Light Change | 0.05 |
| | Probability | 0.75 |
| Affine Transformation | Maximum Wrap | 0.0 |
| | Minimum Scale | 0.8 |
| | Probability | 0.8 |
| Normalisation | ImageNet Statistics | Mean: [0.485, 0.456, 0.406],<br>Std: [0.229, 0.224, 0.225] |
| | Macenko | **Reference image**<br>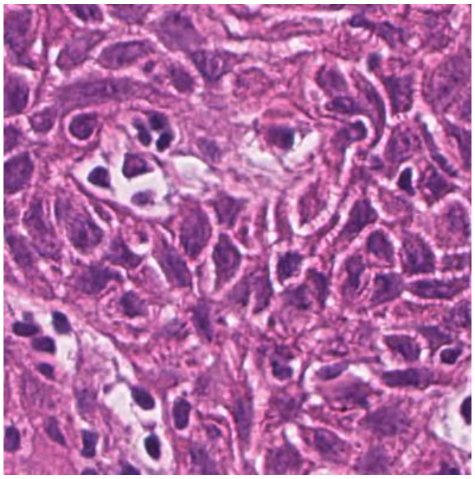 |

**eTable S3** *Performance of the multi-resolution model on validation sets* (NMSCS and HD) for classifying different subtypes of skin cancer, including melanoma and non-melanoma types.

| Class | n | AUROC Mean [95% CI] | Accuracy Mean [95% CI] | Precision Mean [95% CI] | Recall Mean [95% CI] | F1-score Mean [95% CI] |
|---|---|---|---|---|---|---|
| BCC | 3236 | 0.999 [0.999, 0.999] | 0.996 [0.995, 0.996] | 0.937 [0.929, 0.945] | 0.947 [0.939, 0.954] | 0.942 [0.936, 0.948] |
| IEC | 1421 | 0.990 [0.989, 0.991] | 0.988 [0.987, 0.989] | 0.611 [0.586, 0.635] | 0.641 [0.616, 0.665] | 0.625 [0.604, 0.646] |
| Melanoma | 1220 | 0.100 [0.999, 1.000] | 0.999, [0.999, 0.999] | 0.980 [0.972, 0.988] | 0.938 [0.924, 0.951] | 0.959 [0.950, 0.967] |
| Naevi | 944 | 1.000 [1.000, 1.000] | 1.000 [1.000, 1.000] | 0.967 [0.955, 0.978] | 0.998 [0.995, 1.000] | 0.982 [0.976, 0.988] |
| SCC | 7352 | 0.995 [0.994, 0.995] | 0.980 [0.979, 0.981] | 0.896 [0.889, 0.903] | 0.854 [0.846, 0.862] | 0.874 [0.868, 0.880] |
| Non-cancerous | 76380 | 0.998 [0.998, 0.998] | 0.984 [0.983, 0.985] | 0.989 [0.988, 0.990] | 0.992 [0.992, 0.993] | 0.991 [0.990, 0.991] |

Abbreviations: **NMSCS** = Non-Melanoma Skin Cancer Segmentation, **HD** = Heidelberg Dataset, **n** = Number of patches, **AUROC** = Area Under the Receiver Operating Characteristic Curve, **CI** = Confidence Intervals, **BCC** = Basal Cell Carcinoma, **IEC** = Intraepidermal Carcinoma, **SCC** = Squamous Cell Carcinoma.

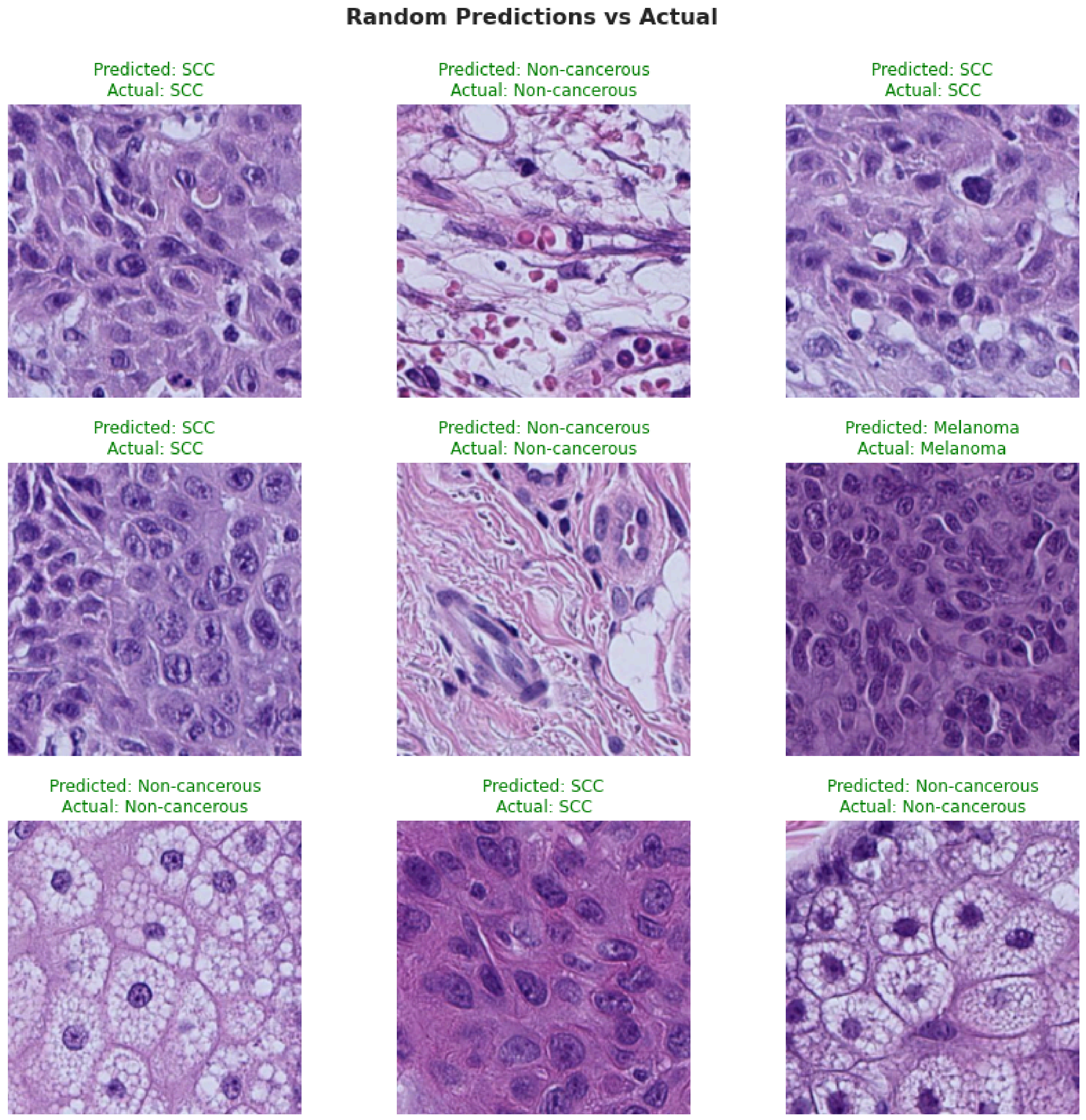


**eFigure S1** *Nine random patches for classifying non-melanoma skin cancer subtypes using a multi-resolution model.*

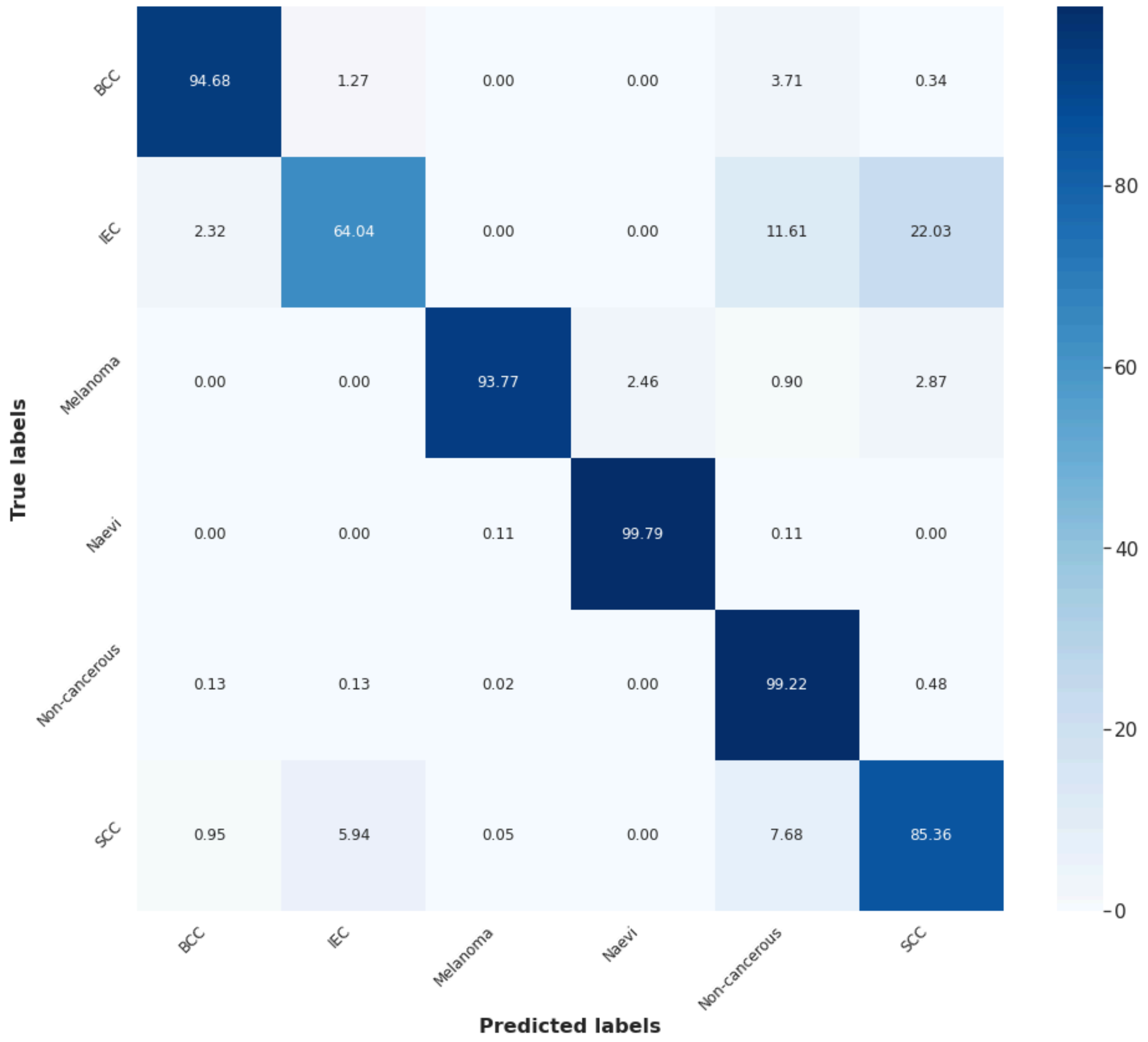


**eFigure S2** *Confusion matrix for six classes on the validation set with 90,553 patches at 10x, 20x, 40x, and 400x magnifications from the Non-Melanoma Skin Cancer Segmentation Dataset and Heidelberg Dataset using the multi-resolution model.*

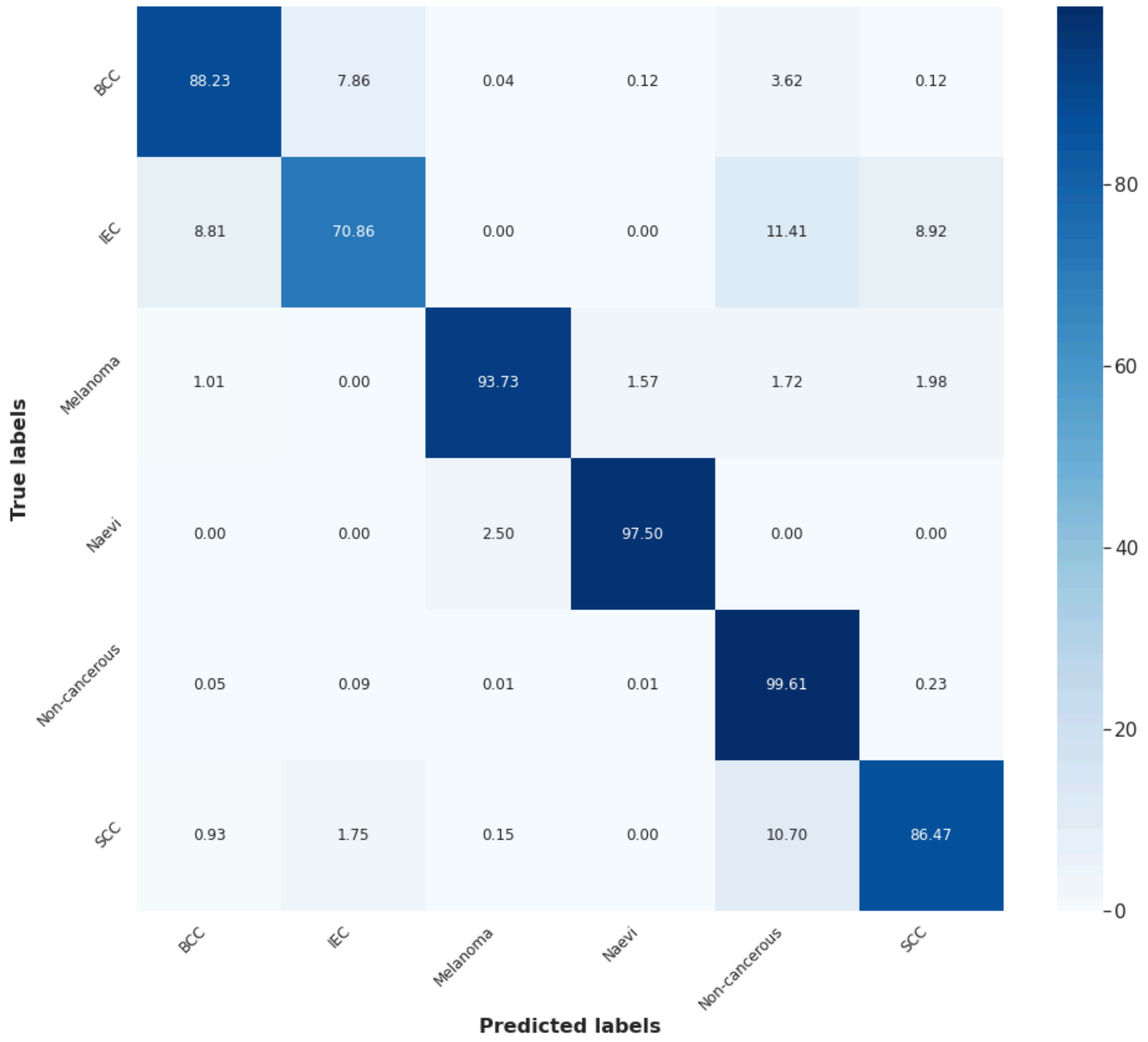


**eFigure S3** *Confusion matrix on the testing set using 10x, 20x, 40x, and 400x patches from Non-Melanoma Skin Cancer Segmentation and Heidelberg Dataset sets, demonstrating the multi-resolution model for classifying six classes efficiently.*

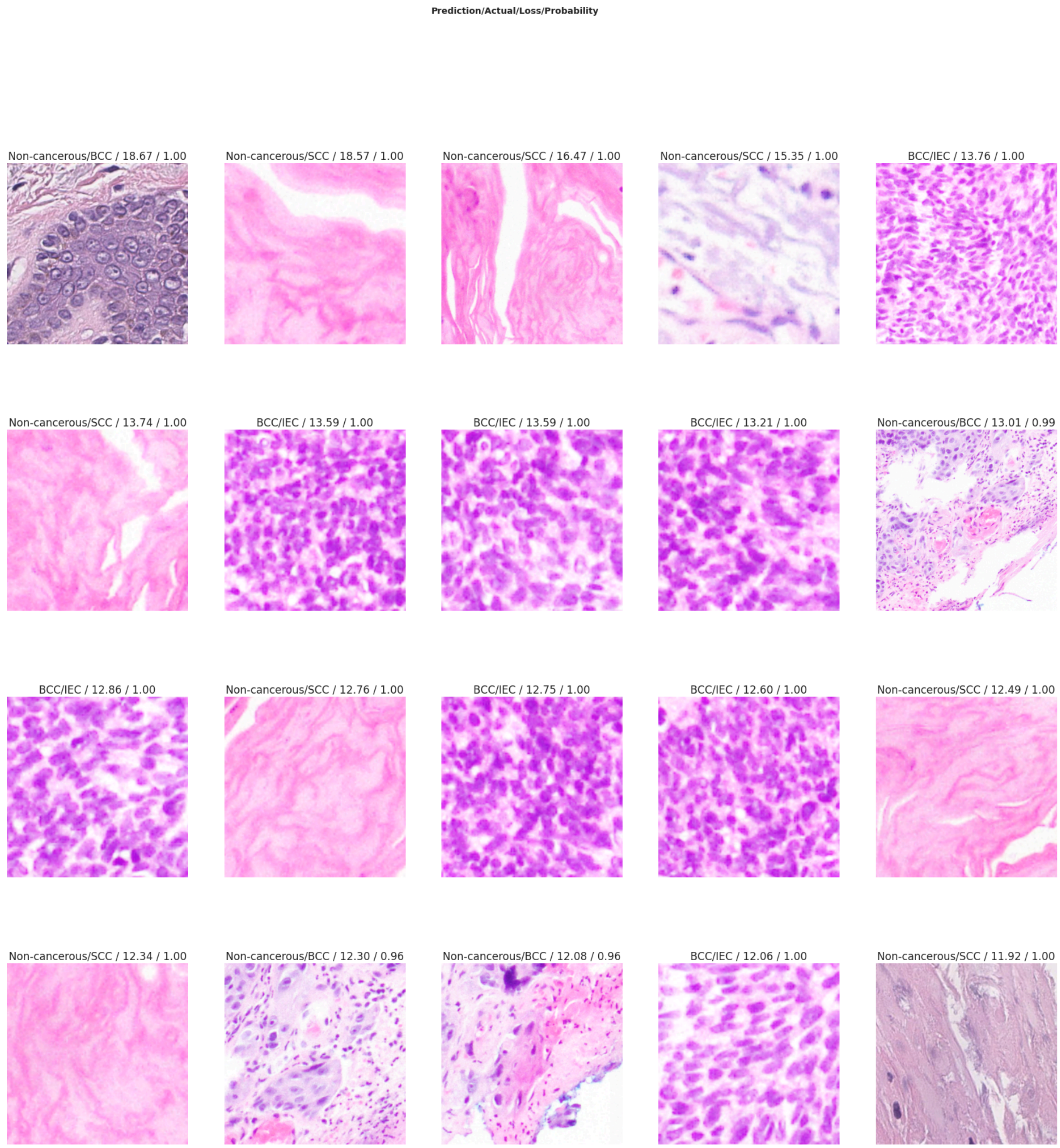


**eFigure S4** *Top 20 misclassified patches were predicted and visualised using the multi-resolution model for six classes on the testing set. These patches represent classes where the model's predictions were incorrect, with a high loss value indicating substantial deviation between the predicted class and the actual label.*

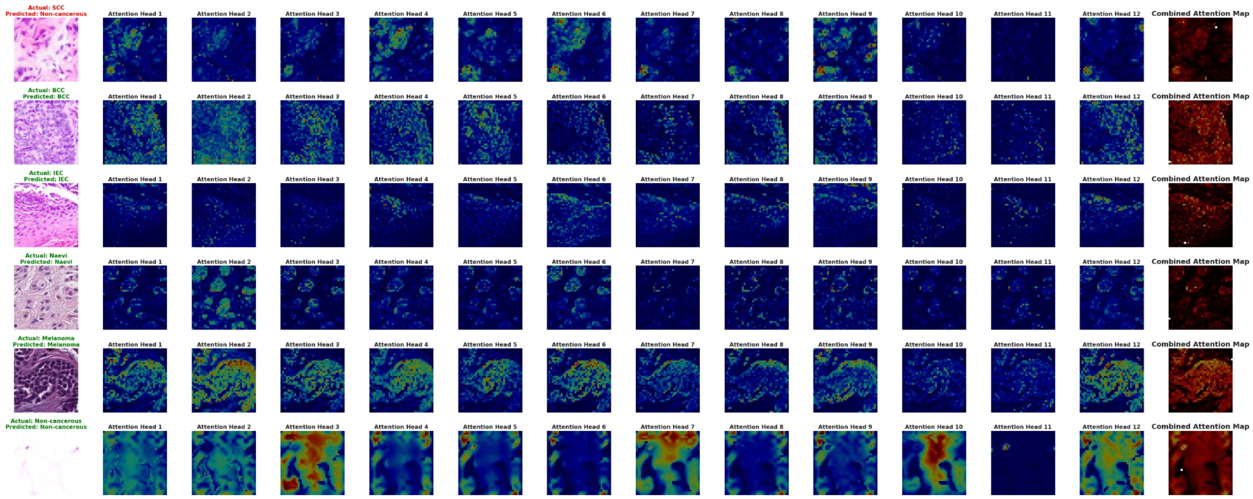


**eFigure S5** *Attention maps from the 12 attention heads of the multi-resolution model for classifying six classes. The first column (left) represents the actual and predicted patch, and the last column (right) represents the combined attention maps.*

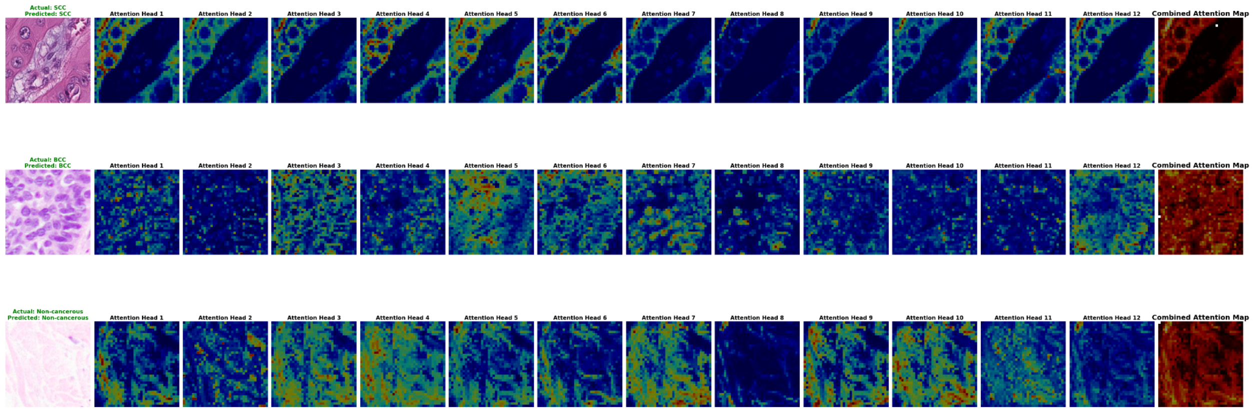


**eFigure S6** *Attention maps from the 12 heads of the multi-resolution model for classifying non-melanoma skin cancer subtypes into three categories. The left column displays the actual and predicted patches, while the right column shows the combined attention maps.*

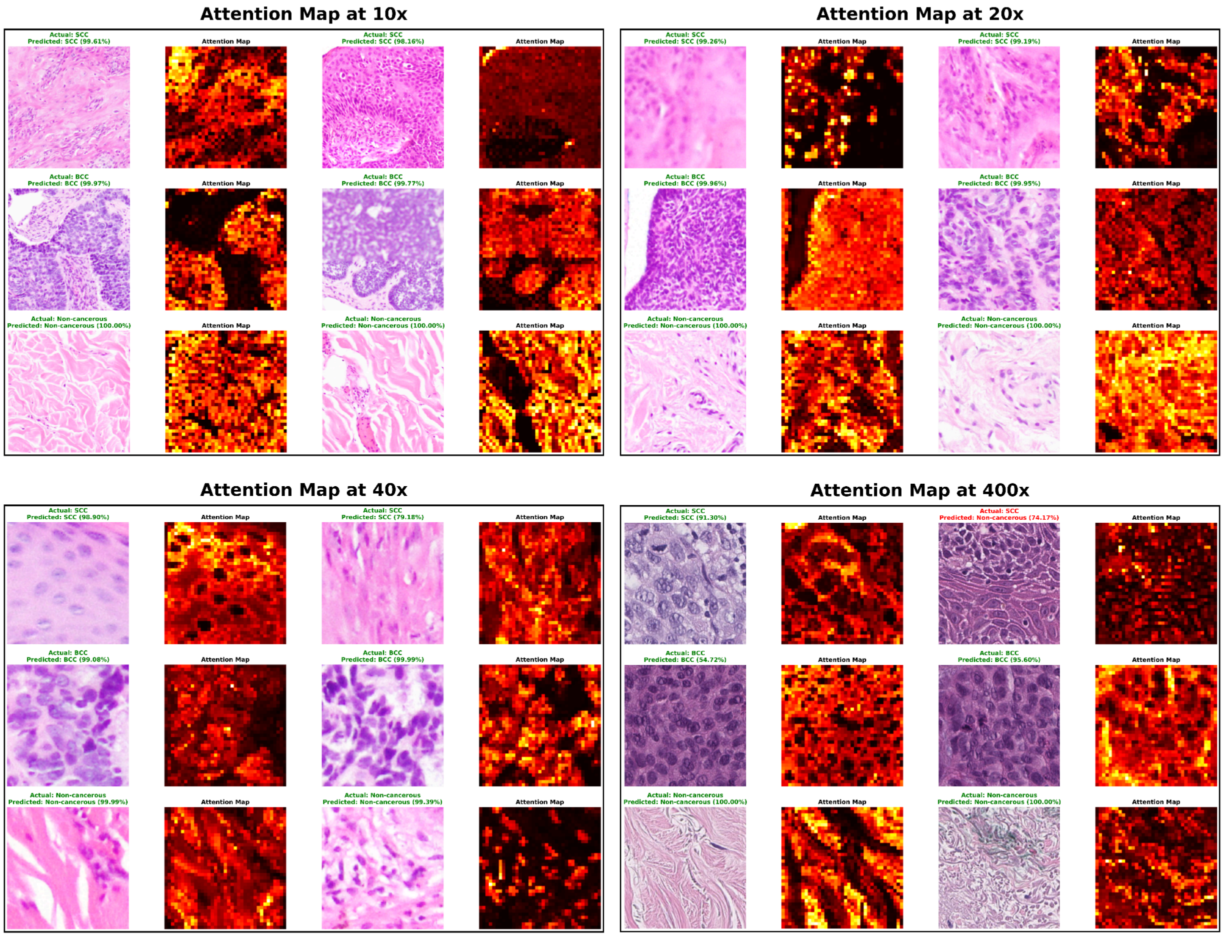


**eFigure S7** *Visualisation of attention maps at different magnifications (10x, 20x, 40x, and 400x) of patches on the testing sets for classifying basal cell carcinoma, squamous cell carcinoma, and Non-Cancerous classes, showing each pair of patches with attention maps, actual class, and predicted class with prediction probability.*
